# Improvement in drug prescription skills in medical students through in-person and remote simulated interviews

**DOI:** 10.1101/2023.04.11.23288429

**Authors:** C Michan Malca, S Christian Siccha, F Ernesto Cardenas, M Maritza Placencia

## Abstract

**Introduction:** Development of drug prescription skills poses critical challenges in medical education. This study determined the effects of simulated interviews on the improvement of drug prescription skills among medical students in 2020.

**Methodology:** This was a quantitative, cross-sectional, analytical, quasi-experimental study of simulated interviews for improving rational drug prescription skills in medical students. Baseline, pre-, and post-intervention assessments of prescription skills were performed using an expert-validated instrument constructed from the WHO Good Prescribing Guide. Three simulated interviews with different simulated patients were conducted in two groups: in-person in the first batch and remotely in the second batch due to mandatory social distancing during the Covid-19 pandemics. Friedman, Dunn-Bonferroni, and Wilcoxon tests were used, considering a significance of level *p*<.05 and standardized mean difference (*Hedges g*); data were analyzed using Excel 2016 and SPSS 28.

**Results:** Fifty-four students completed the required assessments; in-person 28 and remotely 26. The total score for pharmacological prescription skills increased significantly from pre- to post-intervention measurements, from 12.72 +/-2.94 to 15.44 +/-2.50, respectively (*p*<.0001) (*g*: 0.996), and the increase from baseline to post-intervention scores for drug prescription knowledge was 5.39 +/-3.67, 11.28 +/-3.50, respectively (*p* <.01).

**Discussion:** Our results suggest that the implementation of pre-briefing and debriefing strategies in remote and in-person clinical interviews with simulated patients significantly improved drug prescription skills and pharmacological knowledge among medical students. The logical sequence of the WHO Guide for Good Prescribing may have facilitated debriefing, knowledge acquisition, and transfer to various clinical contexts.

## Introduction

Pharmacology learning plays a key role in medical education as it becomes the scientific basis for rational prescription and therapeutics, with a direct impact on patients’ health. However, studies on medical students have found deficiencies in their clinical pharmacology preparation [1] and therapeutic skills [2], possibly because traditional pharmacology courses lack learning experiences immersed in clinical contexts [3]. The earlier students experience educational interventions aimed at developing good prescription skills, the lower the probability that they acquire and perpetuate irrational prescription habits [4].

Many educational interventions have been used to address drug prescription deficiencies with dissimilar findings, including the use of the World Health Organization (WHO) Guide to Good Prescribing [5], an approach with evidence of improvement in pharmacology knowledge, rational prescription, therapeutic skills, motivation, and self-confidence in undergraduate and postgraduate students [6], [7], [8], [9]. Other interventions based on clinical simulation, such as simulated patients, have been found to facilitate knowledge retention, improve drug administration skills, enhance motivation and commitment to learning, and promote patient’s security [10]. High-fidelity scenarios may lead to long-life pharmacological learning [11].

There is a need for more suitable educational approaches with evidence of their positive effects on prescription skills among medical students. We hypothesized that the implementation of simulated clinical interviews methodology may improve rational drug prescription skills in medical students, we started using in-person simulated clinical interviews but due to restrictions on social interactions imposed by the COVID-19 pandemics we were forced to adapt to telesimulation-based remote simulated clinical interviews, posting interesting challenges and accomplishments we report here.

## Materials and Methods

In 2020, a quasi-experimental design was developed to conduct simulated clinical interviews with simulated patients to train medical students to improve their drug prescription skills. Their performance in the simulated interviews was assessed using a rubric based on the steps suggested by the WHO and validated by local experts in pharmacology considering problem identification, therapeutic objective determination, personalized drug selection, drug description, and identification of information for adverse drug reactions (ADRs), based on the steps suggested in the Guide to Good Prescribing from the WHO [5]. We included medical students who had passed a pharmacology course, had no previous experience with simulated patients in pharmacology, and agreed to sign the informed consent sheet. They received information about the drugs that would be used the following week in the simulated clinical interviews and the WHO guide for good prescribing.

Before starting the first simulated interview, students were assessed for their baseline prescription knowledge in a clinical case using an initial short-answer test based on the WHO guidelines. Subsequently, during the simulated interviews, pharmacology teachers trained in simulation methodology assessed students’ prescription skills, and all completed the following assessments: baseline (1), pre-intervention (2), and post-intervention (3). The educational intervention consisted of *pre-briefing, debriefing*, and feedback simulation strategies implemented during interview 2, so that its impact could be assessed on students’ performance during interview 3. All three cases were different (hypertension, ankle pain, asthma), and the students’ performance was assessed by different teachers. Students’ participation ended when they completed the final short-answer test for a different clinical case. The assessments and interventions had to be performed in two groups: in-person at the simulation facility before the lockdown decreed due to the Covid-19 pandemics and remotely using the Zoom® platform, both with the same procedure described. The simulated patients were trained using Debra Nestel’s proposal [12], and the assessment criteria were standardized in two coordination meetings.

Differences in the scores obtained in the skills assessments were analyzed using the Friedman repeated measures test for non-parametric distributions and the Dunn-Bonferroni test to identify significance between paired measurements. Comparisons between in-person and remote interventions were performed using the Mann-Whitney U test. Differences between the results of the knowledge tests were analyzed using the Wilcoxon test for paired measurements. Statistical significance was set at *p* value < 0.05. The standardized Mean Difference (SMD) (*Hedges’ g*) was also calculated. Analyses were performed using SPSS Statistics 28 (IBM, USA) and Microsoft Excel 2016. This study was approved by the Ethics Committee of the Medical School of the National University of San Marcos, Lima, Peru.

## Results

The number of students who agreed to participate in the study was 77; those who completed all the evaluations were 54: 24 men and 30 women (55%), 33 of the participants (61%) were in their third year of study, and 21 in their fourth and fifth years.

The scores obtained for drug prescription skills in the interviews with simulated patients increased from baseline (11.89 ± 3.40) to pre-intervention (12.72 ± 2.94) and post-intervention levels (15.44 ± 2.50) See Figure 01. These repeated measures were analyzed with a non-parametric Friedman test, resulting in a significant *X*_*r*_ ^*2*^=54.78, (*p* < .001); therefore, the null hypothesis was rejected. We completed pairwise comparisons, with statistical significance adjusted using the Dunn-Bonferroni test. We found significant differences between the pre- and post-intervention paired scores (*p*<.0001) (*Hedges’ g*_*s*_: .996); however, no significant differences were found between the baseline and pre-intervention paired scores (*p*=.146). See Figure 02.

**Figure 01.**
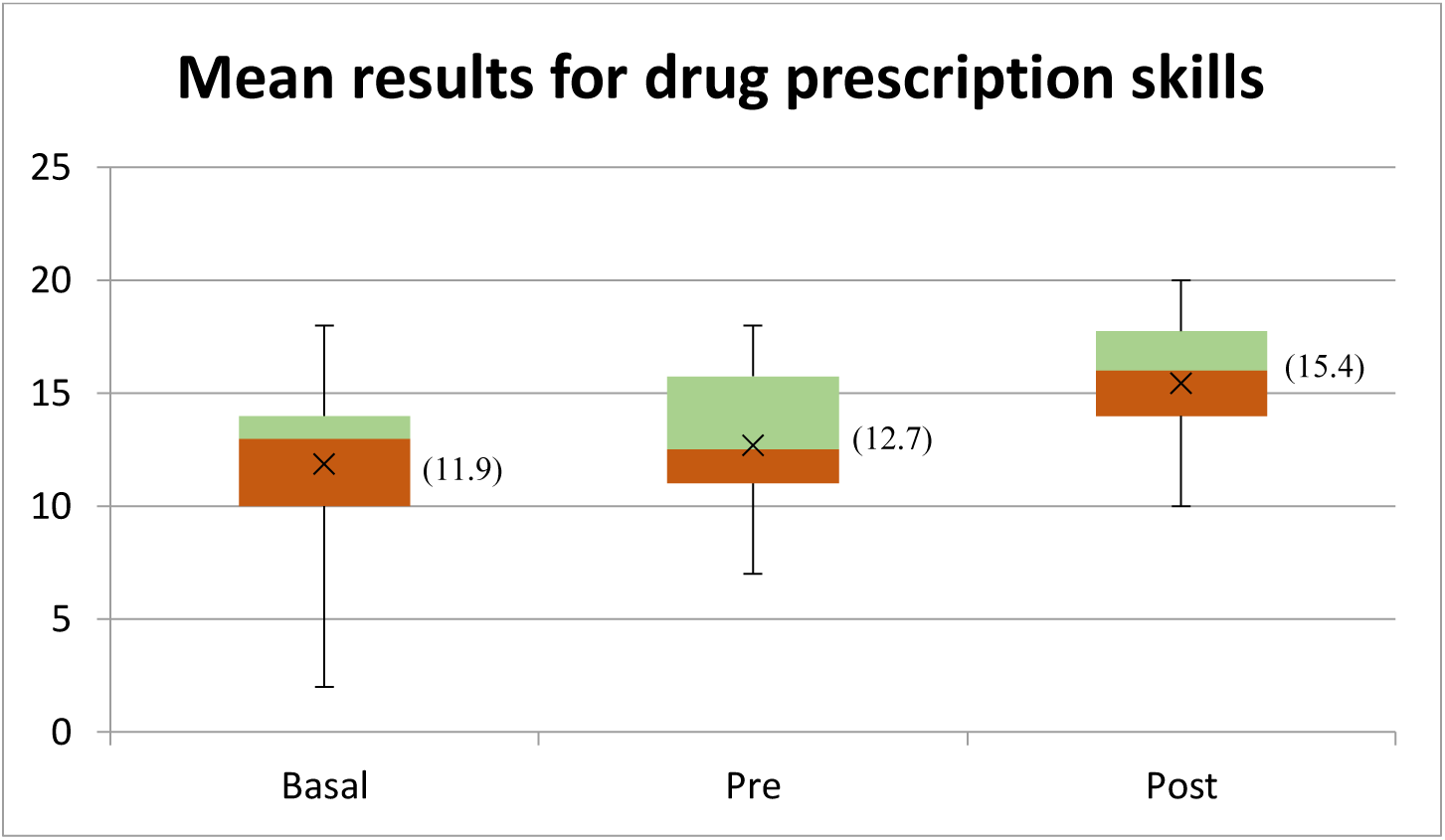
Mean results for drug prescription skills: Baseline, Pre-intervention and Post-Intervention. Own data.

**Figure 02.**
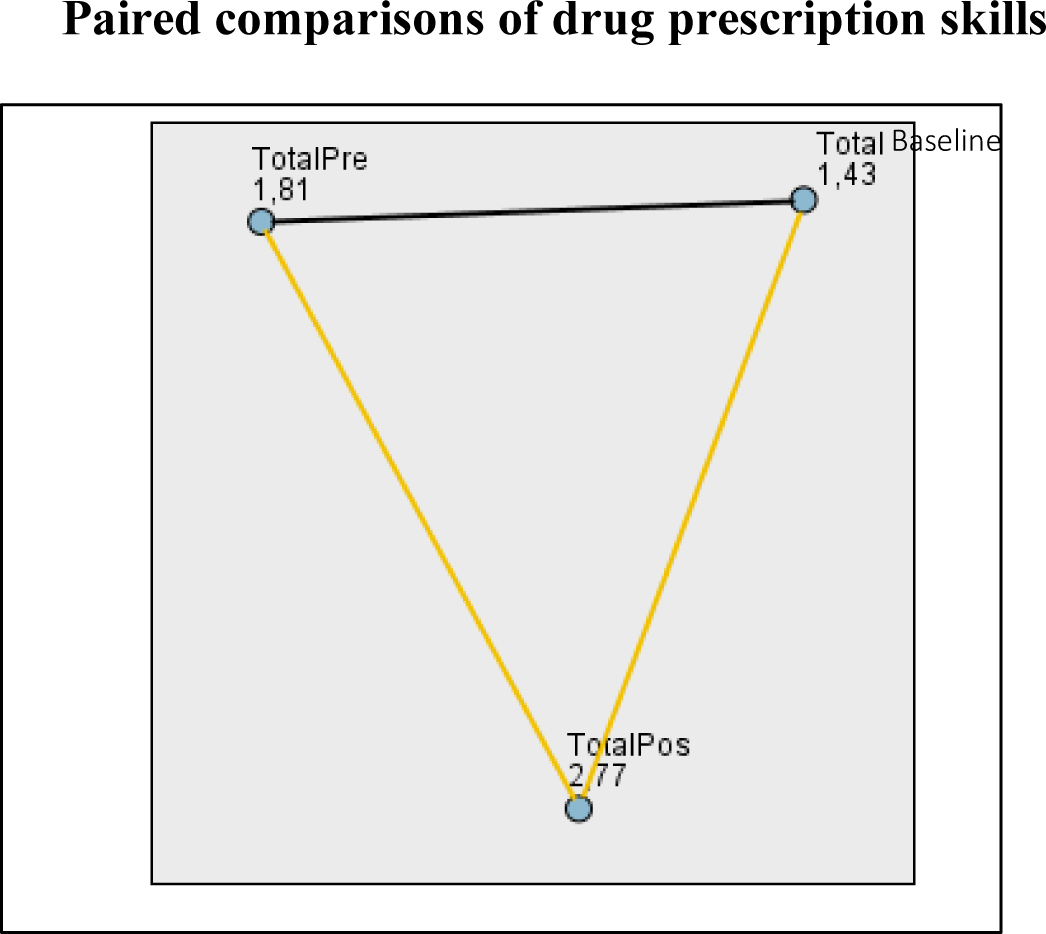
Paired comparison of drug prescription skills: Total-Baseline, Total-Pre-intervention, and Total-Pos-intervention. Yellow lines represent a statistically significant difference and dark lines represent no statistically significant difference. Values represent mean ranks. Own data

The scores obtained for all the components of drug prescription skills (definition of patients’ problems, definition of therapeutic objectives, selection of personalized drug, description of personalized drug, and identification of information for ADRs) in the interviews with simulated patients showed increases, which, when analyzed with non-parametric Friedman tests, resulted in statistically significant differences; therefore, the null hypothesis was rejected. Paired comparisons of pre- and post-intervention scores adjusted using Dunn-Bonferroni tests showed significant differences in all components except for the definition of patients’ problems. The same comparison between the baseline and pre-intervention paired scores showed no significant differences. The details are presented in Table 01.

**Table 01.** Statistical analyses of drug prescription skills.

| | Friedman test for repeated measures<br>$N = 54$ | | $p$ value for paired comparisons with Dunn-Bonferroni test | | |
| --- | --- | --- | --- | --- | --- |
| Variable | Statistic<br>$X_r^2$ | $P$ value | Pre vs Post Intervention | Baseline vs Post intervention | Baseline vs Pre intervention |
| Drug Prescription Skills | 54.78 | < .001 | = .000 | = .000 | = .146 |
| Definition of patient problems | 7.75 | = .021 | = .146 | = .408 | = 1.000 |
| Definition of therapeutic objectives | 26.504 | < .001 | = .014 | = .000 | = .805 |
| Selection of personalized drug | 41.97 | < .001 | = .000 | = .000 | = 1.000 |
| Description of personalized drug | 22.98 | < .001 | = .018 | = .000 | = .447 |
| Definition of information for ADRs | 19.254 | < .001 | = .048 | = .004 | = 1.000 |

Source: Own data

Comparison of the mean scores obtained for drug prescription skills in the in-person and remote group interviews with simulated patients did not show significant differences at baseline (Mann-Whitney test, U = 264.00, *p* = .081); however, they were significantly higher in the remote group at the pre-intervention and post-intervention levels (U = 188.50, *p* = .002) and (U = 139.00, *p* < .001), respectively. In the paired comparison with Dunn-Bonferroni test, significant differences were found between the results of pre- and post-intervention measurements in-person (*p*=.000) (*g*_s_=1.072) and remote groups (*p*=.009) (*g*_s_=1.143), respectively. The results of the comparison of mean scores obtained for all components of drug prescription skills in the in-person and remote simulated interviews indicated that globally, the intervention effect was significant in all except the problem definition component, and the increase was not significant between the baseline and pre-intervention measures. Increases in repeated measures were significant, except for the definition of patient problems and description of personalized drugs in the in-person group.

The increase in the test of cognitive abilities from baseline to post-intervention, was further analyzed using a Wilcoxon signed-rank test, which indicated that in the in-person and remote groups, the post-intervention scores were statistically higher, (Z = -4.628, *p* <.001) and (Z = - 3.814, *p* <.001) (*g*_s_=1.642). The Mann-Whitney tests for mean scores of cognitive abilities for drug prescription between the in-person and remote groups showed non-significant differences at baseline and significant differences at the post-intervention level (U = 263,500, *p* = .080) and (U = 133,000, *p* < .001, respectively).

## Discussion

In the present study, our results showed significant improvements in medical students’ drug prescription skills after the implementation of remote and in-person simulated clinical interviews with simulated patients. Their performance was assessed by trained teachers using an instrument based on the Guide to Good Prescribing by the WHO. The improvement was significant only when the students experienced all phases of the planned intervention: pre-*briefing*, simulated encounter, and *debriefing*, as evidenced by the significant increases in the scores from the pre-intervention to post-intervention measurements. The non-significant increase in the comparison of results from baseline to pre-intervention, when they experienced only the simulated interview, is consistent with what has been observed in other studies on the importance of pre-*briefing* and *debriefing* in achieving the best learning results with the use of simulation. The debriefing in our research was carried out after the simulated encounter, considering the WHO Guide to Good Prescribing, including the following stages: reaction/description of the event, understanding/analysis of what happened in the simulation, and application/summary of what has been learned using the Plus-Delta model, suggested by various authors to achieve greater efficiency, emphasizing students’ self-assessment [(13)]; [(14)]; [(15)]; [(16)]. Two other studies have highlighted the importance of the reflective process of debriefing, both in person and virtual, as a fundamental element for a clear understanding of the actions and cognitive processes that promote learning and clinical performance in the future [(17)]; [(18)].

The implementation of the WHO Guide for Good Drug Prescribing as the basis for developing simulated interviews and debriefing likely had a positive impact on the improvement of drug prescription skills among the participants in this study. This guide allowed the application of a reasoned process for selecting drugs according to the identified problem and the proposed therapeutic objectives, consistent with what was reported by de Vries et al., who demonstrated a positive effect for retention as well as for knowledge transfer, and concluded that the WHO 6-step method has the greatest amount of supporting evidence as a strategy to improve rational prescribing, [(19)] which supports the results obtained in this investigation. Likely, a study in Turkey found a beneficial impact on rational prescribing skills in 4th year medical students in the short term with a course of rational pharmacotherapy, measured through a pre- and post-intervention OSCE, which was based on the six steps of the WHO Guide for Good Prescribing, and the students also expressed satisfaction with the course [(20)].

Our results are also supported by the varied evidence on the use of simulation methodology and the modality of simulated or standardized patients for learning pharmacology in different contexts, which demonstrate the feasibility of developing simulation experiences and its positive effects on learning to prescribe medicines. In a study conducted in Turkey similar to the present investigation, the results of the post-test demonstrated effectiveness with the use of this type of simulation [(21)]. In Japan and the USA, there is evidence of the effectiveness of the use of simulated patients in improving communication skills, safety in medication administration, and confidence in nursing and pharmacy students, showing the same effectiveness as traditional pharmacology classes [(22)]; [(23)]; [(24)]; [(25)].

Additionally, the learning strategy used in this research agrees with that suggested by British authors on teaching safe prescriptions to medical students, which requires the contextualization of learning with a patient in a properly evaluated participatory process, allowing the application of knowledge in the management of a patient, such as in a simulated prescribing practice that requires appropriate knowledge, skills, and behaviors [(26)]. In the case of prescription skills, such as those studied in this work, learning activities must include decision making and communication of the prescription in writing or electronically [(27)].

The increase in knowledge indicated for pharmacological prescriptions agreed with improvements in knowledge and skills found in a 2016 study [(28)]. Although there was no formal randomization, notably the in-person group obtained significantly higher means in the post-intervention knowledge measurement than the remote group, which could be due to the lack of experience with remote learning in pharmacology and the reduced experience of medical students in developing small paragraph response exams in the remote group.

In this study, each simulated interview was assigned with a different clinical case, with participants obtaining significant improvements in their drug prescription scores. Hence, it is likely that this strategy will help transfer theoretical and practical knowledge of pharmacology to a feasible clinical learning context for rational drug prescription. This possible transfer effect had previously been suggested in an evaluation of the effect of a preclinical pharmacotherapy program on the quality of rational prescribing by medical students during rotations in internal medicine, which found greater use of rational prescribing in trained topics and in other clinical situations, which implies a transfer effect by the program [(29)]. This finding may be a starting point for developing the strategy since studies report gaps in knowledge about transfer, scalability, cost-benefit, and the most appropriate instructional design to implement the strategy of simulated or standardized patients in pharmacology education for health science students [(30)], [(31)].

In the intergroup analysis, our results suggest that regardless of the intervention group, no differences were found in the statistical significance of the results for drug prescription skills and the personalized drug selection, which is the most important component of the WHO Guide. This is a key finding, because for selection of the personalized drug, its efficacy and safety must be considered in the clinical and physiological contexts of the patient, as the integration and application of what has been learned in pharmacology courses [(5)].

Possible reasons for the greater effect in the remote group would be more motivation, greater comfort (higher psychological safety), greater self-regulation, and better achievement of knowledge and skill goals. In the absence of an in-person instructor, students probably assume greater commitment to their learning, as reported in various publications [(32)]; [(33)]; [(34)]. In a comparative qualitative study of three modalities of simulation and feedback in health education, they found that simulation with actors allowed the consolidation of knowledge and learning, the practice of communication skills, and a high level of interactivity, but that it could generate anxiety and nervousness in students with the pressure of interview time limiting its effectiveness [(35)], which may help explain some of the heterogeneous results found in this investigation.

The present investigation used the Zoom® platform to develop the remotely simulated interviews, an example of telesimulation. The challenges generated by social distancing due to the Covid-19 pandemic have driven the adoption of innovations and the awareness of connectivity limitations in our homes and institutions, as well as the need for contingency plans based on circumstances and available resources [(36)]. This study represents the implementation of an alternative solution to this health crisis context. However, further analysis of the applications of remote teaching and learning in pharmacology is required [(37)]. A study in Romania highlighted that the pandemic has generated new approaches to learning and that the benefits of remote education should be maximized, proposing hybrid approaches to take advantage of both in-person and remote education systems [(38)]. In another aspect to be investigated, it is asserted that there is evidence that prescription behaviors are solidified in medical schools and that the cause of prescription errors could be attributed to the lack of integration of scientific knowledge with clinical experience; however, there is room for improvement in medical education in pharmacology [(39)].

The changes in healthcare professionals’ education have crafted new teaching paradigms, which should be characterized by more integrated and contextualized learning experiences, development of competencies, formative and summative assessments with adequate feedback, and faculty training accordingly [(40)]. By 2017, only 4% to 24% of the pharmacology teaching programs had used simulation to train patient safety and in problems related to medications, which represents great potential for innovation in strategies and tools in the pharmacology teaching-learning process. The use of simulated patients, incorporating a positive emotional charge with an active, student-centered approach, may help them better understand and retain more information for a long time [(36)]. Medical and nursing students accept the use of simulated or high-fidelity patients before contact with real patients, with high levels of satisfaction due to the safe and authentic environment, as well as the greater ease of applying knowledge and skills in interprofessional and interdisciplinary education [(41)].

The main limitation of this study was that the sample of participants was not complete due to mandatory social distancing, which changed the priorities of the students for their collaboration with the research, but it opened the possibility of studying the simulation strategy remotely. The strategy of selecting participants for convenience limits the generalizability and implications of the results. There was no randomization to remote or in-person groups, but this strategy may be used for future research in telesimulation. The adaptation of the in-person procedure to virtuality highlighted the difficulties in non-verbal communication and the impossibility of examining the patient, as well as the lack of preparation of the participating students to conduct interviews with remote simulated patients, which was carried out partially in the present investigation within the pre-briefing and debriefing processes. The selection of cases had to be limited to situations with a low level of complexity, where the information required to define the patient’s problems had to be obtained only from the interview; consequently, the range of clinical situations was limited. It was not possible to differentiate between the effects attributable to the simulation methodology or the WHO guide for good prescription in the improvements found in this study, which may be summative or synergistic, an interesting topic for future research.

The implementation of remote and in-person simulated clinical interviews with simulated patients had a positive effect on the development of rational drug prescription and cognitive skills in medical students. The integration of the WHO Guide for Good Prescribing facilitated the debriefing phase of the intervention and knowledge acquisition thanks to the logical sequence of its steps. The telesimulation strategy can be used to train health professionals to develop communication, decision making, and clinical reasoning skills. The improvement in prescription skills using different cases in simulated interviews may represent the transfer of theoretical knowledge to varied clinical contexts for learning pharmacology, with resources easily available to medical schools.

## Data Availability

All data produced in the present study are available upon reasonable request to the authors

## Acknowledgments

Special thanks to the simulated patients Carmen Puma, Anabell Quispe, Rubí Cáceres; assessors Dr. Eleazar Aliaga, Dr. Edgardo Huarhua, Dr. Conrad Ortiz, Dr. Henry Montellanos, Dr Alexis Loza; Dr. Cesar Gutierrez, Dr. Paolo Wong, and all participants.

